# Pelvic floor health in athletics: a cross-sectional study at the Lima World Athletics U20 Championships

**DOI:** 10.1101/2024.08.24.24312375

**Authors:** Giagio Silvia, Bermon Stephane, Garrandes Frederic, Rial Rebullido Tamara, Pillastrini Paolo, Vecchiato Marco, Adami Paolo Emilio

## Abstract

**Objectives:** This study aims to investigate various aspects of pelvic floor health among male and female athletes participating in the Lima World Athletics U20 Championships.

**Methods:** This observational, cross-sectional study will be conducted through a web-based survey via SurveyMonkey. The survey will collect demographic and anthropometric data, as well as medical history, including any injuries to the lower abdomen or pelvic area and history of urinary tract infections. It will also gather information on athletics-related characteristics, such as event specialization, training intensity, and competition volume. Additionally, the survey will explore pelvic floor health by assessing athletes’ knowledge of pelvic floor function and dysfunctions, as well as related behaviors. The prevalence rates of pelvic floor dysfunction (PFD) —including urinary and anal incontinence, pelvic pain, overactive bladder, and pelvic organ prolapse—will be assessed, along with their impact on sports participation and potential risk factors. Urinary incontinence will be further evaluated using the International Consultation on Incontinence Questionnaire-UI Short-Form (ICIQ-UI-SF), with a focus on identifying specific triggers related to athletic activities. Participants will also provide information about their experiences with pelvic floor screening, discussions about these issues, symptom management strategies, and any medical assessments or treatments they have received. Data will be analyzed using descriptive statistics and subgroup analyses. Correlation between collected variables and the presence of PFD will be investigated using correlation analyses.

**Conclusion:** The findings from this study will provide valuable insights into the unique challenges faced by young elite athletes concerning pelvic floor health. This research represents an initial step toward an international initiative, promoted by World Athletics, to enhance pelvic floor health surveillance and promote overall well-being in athletics.

## INTRODUCTION

According to literature, both male and female athletes engaging in sports may experience various pelvic floor dysfunction (PFD) [1–3], including urinary incontinence (UI) [4–11]. High-impact sports, like athletics, have been linked with an increased risk of developing PFD due to the repetitive ground impacts and increased intra-abdominal pressure, which place significant stress on and train the pelvic floor muscles [10,12–15].

Despite the increasing recognition of pelvic floor health’s importance in sports medicine and the growing interest in this research topic [9,11,16–19], existing studies mainly focused on female adult athletes with UI, highlighting a research gap concerning high-level young athletes [20] of both sexes with multiple PFD symptoms [6]. Additionally, only a few authors investigated these symptoms among athletes participating in athletics [15,21–23].

In light of this context, the main aim of this study will be to comprehensively investigate different aspect of pelvic floor health among male and female athletes participating in the Lima World Athletics U20 Championships scheduled for 27-31 August 2024. Specifically, this study will aim to assess PFD prevalence rates, their impact on sports participation, and to identify potential associated risk factors. Our assessment will encompass a range of PFD, including UI, anal incontinence (AI), pelvic pain, overactive bladder (OAB) symptoms, and pelvic organ prolapse (POP). Females’ urogynaecology domains will be further assessed. Additionally, the research will aim to explore athletes’ awareness, knowledge, and behaviours related to pelvic floor health.

By addressing these gaps in the literature, findings will provide valuable insights and information into the unique challenges faced by young elite athletes. This study represents the starting point for an international initiative promoted by World Athletics aimed at promoting pelvic floor health surveillance and overall well-being in athletics.

## METHODS

An international research team work on the present observational, cross-sectional study, through a web-based survey. The study has received approval from the Bioethics Committee of the University of Bologna, Italy (Prot. N. 0127854, 08/05/2024) and will be conducted in strict accordance with the Declaration of Helsinki. Informed consent will be obtained from all study participants. For the reporting, The Strengthening the Reporting of Observational Studies in Epidemiology (STROBE) [24] and the Checklist for Reporting Results of Internet E-Surveys (CHERRIES) guidelines [25]will be used. This research project is supported and promoted by World Athletics.

### Research team

The author team comprises two women and five men, including clinicians and researchers from various specialties and nationalities representing Italy, Spain, France and the USA, The team expertise includes the following: sports medicine, musculoskeletal/sport physiotherapy, pelvic floor physiotherapy, urogynaecology and exercise physiology.

### Data collection

Target research population will involve male and female athletes eligible for and participating in the Lima World Athletics U20 Championships. Participation is voluntary, and a gadget will be offer as incentive. An electronic educational leaflet containing information on pelvic floor health will be provided to all athletes upon completion of the survey.

#### Procedure

Before the Championships, all 214 members of the World Athletics federations will receive a briefing via email outlining the nature and goals of the survey. They will be requested to disseminate this information to their respective teams one week before the Championships and obtain informed consent. Upon arrival at the event, registered athletes will be encouraged to visit Health & Science (H&S) booths located in the competition area. Additional booths will be placed in the hotel entrances. World Athletics medical staff (PEA, FG, SG) will be available to answer questions, provide medical counselling, and assist athletes in filling out questionnaires. At these sites, personal accreditation badges will be scanned to confirm identity and prevent biases. Athletes meeting these criteria will receive an invitation pamphlet containing the QR code with direct link to the survey page.

#### Survey

Data will be collected via an online questionnaire using SurveyMonkey software. Participants will be able to review and check the completeness of the survey and eventually change responses using a back button, before submitting their answers. Data will be then downloaded and stored in an encrypted computer, and only the first author (SG) will be access to the information during all stages of the study. Since all personal data will be collected anonymously, participants will be ensured that their identities would not be disclosed.

#### Question domains and questionnaires

The survey will consist of four domains and take approximately 10 min to complete. The selection of questions and questionnaires considered the athletes’ age and their ability to comprehend medical textual material. The survey was piloted with a small group of athletes and with the research team to assess the feasibility, usability, and clarity of the survey, reviewing the questions and giving feedback on the content. At the beginning of the survey, athletes will have the option to choose from English, French, Spanish and Italian languages. Questions will include the following domains:

1. Demographics and anthropometric measures (e.g., age, sex, BMI, geographical area),
2. Medical history e.g., muscle or bone injuries in your lower belly or pelvic area, frequent urinary infections),
3. Athletics-related characteristics (e.g., event specialization, training and competing volume),
4. Pelvic floor health (e.g., awareness and knowledge of pelvic floor and PFD, pelvic floor behaviours). This section also will ascertain prevalence rates of PFD in both sexes. In particular, presence of UI will be in deep investigated using The International Consultation on Incontinence Questionnaire-UI Short-Form (ICIQ-UI-SF), patient reported outcome measures with grade A recommendation from the International Consultation on Incontinence (ICI) [26]. Presence of athletics-related UI will be further investigated. Particularly, athletes who will report athletics-related UI will be asked what triggers (e.g. event specific training, competition, or other related activities) cause symptom and its impact on participation.

To assess prevalence rates of other PFD, ad hoc questions will develop accordingly to population-based study in previous studies [10,27,28]. This choice is underpinned by three key considerations. Firstly, there is a notable absence of a comprehensive tools tailored to assess PFD symptoms altogether. Secondly, utilizing questionnaires containing numerous irrelevant items could potentially deter participation, as evidenced in previous studies [27,29]. Thirdly, in alignment with the study’s objectives, our survey will not aim to establish medical diagnoses. Instead, questions about PFD symptoms will be extracted from the PFD-SENTINEL tool [30]. This cluster of symptoms has been extracted from validated questionnaires and has also received agreement from an international panel of experts, ensuring a targeted investigation into relevant symptoms prevalent among the athlete population.

Female health will be additional assessed with gynaecological questions for example regarding menstrual cycle and the use of hormonal medications or other contraceptive methods.

All athletes will then be asked about their previous experiences in PF screening, engaging in open discussions with individuals regarding the issue, strategies to mitigate PF symptoms, and any medical assessments and/or treatments.

## STATISTICAL ANALYSIS

After collecting all responses In SurveyMonkey, data will be downloaded and will undergo thorough cleaning and verification to ensure accuracy. Statistical analyses will be conducted using SPSS statistical software package version 24 (SPSS Inc., Chicago, IL). Descriptive statistics (numbers with percentages, mean with standard deviations or median and interquartile range) will be used to report collected variables. Descriptive subgroup analysis will be performed, according to sex, event specialization, and other relevant characteristics. Correlation between collected variables and the presence of PFD will be investigated using correlation analyses.

## CONCLUSION

This research protocol outlines a comprehensive investigation into pelvic floor health among athletes participating in the Lima World Athletics U20 Championships. By assessing level of knowledge, symptoms prevalence rates, and impact on sports participation, this study aims to contribute valuable insights into promoting pelvic floor health and well-being among athletes.

## Data Availability

All data available are contained in the manuscript.

## Acknowledgements

Not applicable.

